# Linking cohort data and Welsh routine health records to investigate children at risk of delayed primary vaccination

**DOI:** 10.1101/2022.04.04.22273336

**Authors:** Suzanne Walton, Mario Cortina-Borja, Carol Dezateux, Lucy J Griffiths, Karen Tingay, Ashley Akbari, Amrita Bandyopadhyay, Ronan A Lyons, Richard Roberts, Helen Bedford

## Abstract

**Background:** Delayed primary vaccination is one of the strongest predictors of subsequent incomplete immunisation. Identifying children at risk of such delay may enable targeting of interventions, thus decreasing vaccine preventable illness.

**Objectives:** To explore socio-demographic factors associated with delayed receipt of the Diphtheria, Tetanus and Pertussis (DTP) vaccine.

**Methods:** We included 1,782 children, born between 2000 and 2001, participating in the Millennium Cohort Study (MCS) and resident in Wales, whose parents gave consent for linkage to National Community Child Health Database records at the age seven years contact. We examined child, maternal, family and area characteristics associated with delayed receipt of the first dose of the DTP vaccine.

**Results:** 98.6% received the first dose of DTP. The majority, 79.6% (*n=*1,429) received it on time (between 8 and 12 weeks of age), 14.2% (*n=*251) received it early (prior to 8 weeks of age) and 4.8% (*n=*79) were delayed (after 12 weeks of age); 1.4% (*n*=23) never received it. Delayed primary vaccination was more likely among children with older natural siblings (risk ratio 3.82, 95% confidence interval (1.97, 7.38)), children admitted to special/intensive care (3.15, (1.65, 5.99)), those whose birth weight was >4Kg (2.02, (1.09, 3.73)) and boys (1.53, (1.01, 2.31)). There was a reduced risk of delayed vaccination with increasing maternal age (0.73, (0.53, 1.00) per 5 year increase) and for babies born to graduate mothers (0.27, (0.08, 0.90)).

**Conclusions:** Although the majority of infants were vaccinated in a timely manner, identification of infants at increased risk of early or delayed vaccination will enable targeting of interventions to facilitate timely immunisation. This is to our knowledge the first study exploring individual level socio-demographic factors associated with delayed primary vaccination in the UK and demonstrates the benefits of linking cohort data to routinely-collected child health data.

## INTRODUCTION

Delay in receiving scheduled vaccinations not only leaves individual children vulnerable to vaccine-preventable diseases but may also compromise herd immunity. Delayed primary vaccination has been found to be one of the strongest predictors of subsequent incomplete immunisation (1, 2). In the USA, infants who received the first dose of Diphtheria, Tetanus and Pertussis (DTP) vaccine on time were twice as likely to be up-to-date by two years of age; those not initiating their immunisations on time were two to three times more likely to be delayed for other immunisations given prior to two years of age (3). Children who remain unimmunised or incompletely immunised with primary vaccines are also more likely not to receive MMR (4) or the pre-school boosters (5, 6). Identifying and addressing factors associated with delayed primary vaccination may improve vaccine timeliness, as well as overall vaccine uptake.

Although patterns of vaccination timeliness have been investigated, less research has focussed on factors associated with late receipt of vaccination. Factors associated with adherence to vaccination schedules are complex and context specific (7) and can be different for delayed compared with incomplete vaccination (8). Delayed vaccination has been associated with factors indicative of lower socioeconomic status (8-13), race and ethnicity (8), indigenous ethnic status with geographical remoteness (12), residential mobility and medical conditions in the child (11). In Scotland, using a population database containing immunisation records for over one million children, Friederichs found late MMR vaccination to be associated with deprivation, while the most affluent tended to be vaccinated promptly, or not at all (14). Also, in Scotland, Haider et al (13) found associations between deprivation, uptake and timeliness for four childhood vaccines. Using data for nine London health service areas from the Child Health Information System, Tiley et al (15) reported that although the overwhelming majority of children were vaccinated on time, this was less likely for some ethnic minority groups.

There is limited information about other factors associated with delayed primary vaccination in a UK setting needed to facilitate targeting of interventions.

Delayed vaccinations may suggest broader lack of engagement with health services. In the USA, Rodewald found that among disadvantaged populations, delay in receiving vaccinations was associated with low uptake of other preventive health care services (16). If this were also the case in the UK, then identification and targeting of those who are likely to delay vaccinations may also increase attendance for other child health promotion activities such as routine health and development reviews offered as part of the Healthy Child Programme (17).

This study builds on our earlier research describing the uptake and timeliness of vaccinations amongst Millennium Cohort Study (MCS) participants living in Wales (18). The rich socio-demographic data gathered in the MCS, linked to electronic vaccination records provides a valuable opportunity to characterise children at risk of delayed vaccination. This will facilitate improved targeting of vaccination services. This cohort of children received separate DTP, polio, and *Haemophilus influenzae* type b (Hib) vaccines rather than the pentavalent vaccine (DTaP/IPV/Hib) introduced in 2004. We focus on the DTP vaccine as it was closest to the hexavalent vaccine (DTaP/IPV/Hib/HepB) used currently in the UK. This study explored socio-demographic factors associated with delayed receipt of the DTP vaccine.

## METHODS

### Study population

We used a subset of data from the MCS, a UK-wide nationally representative birth cohort comprising 18,818 children from 18,552 families born between September 2000 and January 2002. Parents were interviewed at home when their child was aged nine months and subsequently at three, five, seven, eleven and fourteen years of age. At the age seven home visit, written consent was sought from parents to link MCS information collected until each child’s 14^th^ birthday, to data routinely collected by government departments or agencies, and other public sector organisations. The Northern and Yorkshire Research Ethics Committee approved the MCS age seven survey; no additional approval was needed for this linked data analysis, which focusses on those resident in Wales. Parents of 1,840 (94.3%) of 1,951 singletons resident in Wales, consented to health record linkage. Linked MCS and National Community Child Health Database (NCCHD) records were available for 1,831 children. We excluded 46 children interviewed in countries other than Wales on one or more occasions by age eleven years and three for whom the main respondent was not the natural mother at the first interview, leaving a final sample of 1,782.

### Record Linkage

We accessed coded data from the NCCHD, which brings together data from local child health system databases held by NHS organisations and includes information from birth registrations, child health examinations and vaccinations.

We used the privacy-protecting trusted research environment in Wales known as the Secure Anonymised Information Linkage (SAIL) Databank to store and access our data. Datasets imported into SAIL are anonymised and linked using a split file process preventing access to both identifiable data and clinical information at the same time. Records are linked through assigning unique encrypted Anonymised Linkage Fields (ALF) to person-based records (19). Additional information on linkage and linkage rates for this project has been published elsewhere (20).

### Timeliness of vaccination

Children born in Wales between August 2000 and November 2001 should have received the DTP vaccine at 8, 12 and 16 weeks of age and a booster dose between the age of three years and four months and five years. Consistent with our earlier methodology (18), the first dose of the DTP vaccine was considered to be delayed if it was received when the child was over 12 weeks of age.

To reduce deductive disclosure, each child’s date of birth was supplied by SAIL as the week of birth (set to the Monday), and a day of birth within that week was assigned by adding a uniform random number between 0 and 6 days. Age at vaccination was calculated using this assigned date and the actual date of vaccination from NCCHD records.

### Potential factors associated with delayed vaccination

As little is known about socio-demographic factors associated with delayed primary vaccinations in the UK, we explored factors associated with wider vaccination outcomes, such as uptake or delay, reported both in the UK and internationally (5-8, 21-29). These included a range of child, maternal, family and area characteristics obtained from MCS sweep 1 data (Table 1).

**Table 1.**
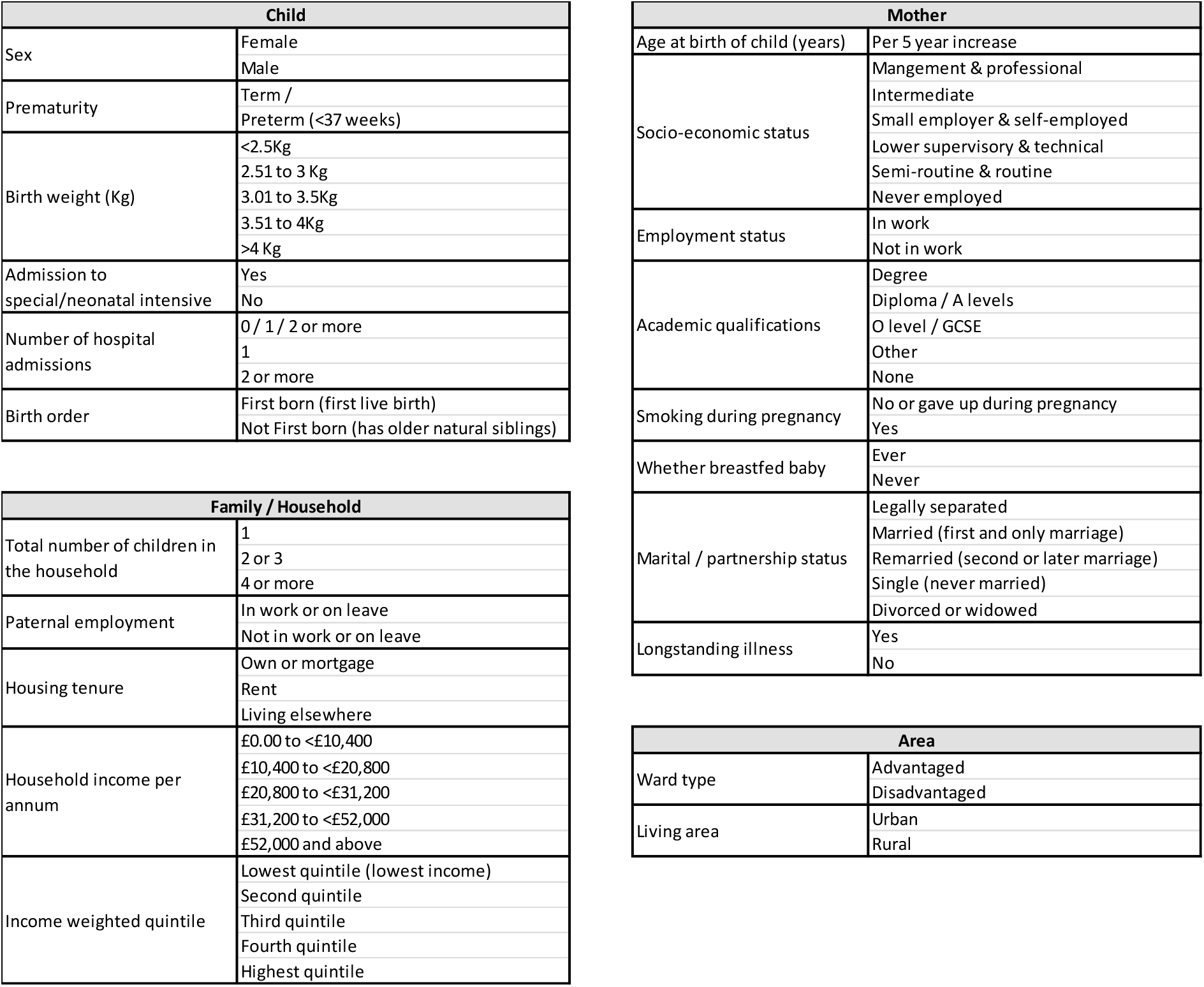
Variables included in the analysis

### Statistical Methods

All analyses were performed using StataSE 13 (StataCorp. 2013. Stata Statistical Software: Release 13. College Station, TX: StataCorp LP.) Survey and non-response weights at age seven years were used to adjust for clustered sampling design, data missing due to losses to follow-up, and lack of consent to linkage (30, 31). Weighted percentages were calculated and reported.

As overdispersion was not present, Poisson Regression models were fitted to explore associations with delayed receipt of DTP. As the outcome of interest was delayed vaccination, those (*n*=23) who had no record of being vaccinated with DTP (up to fourteen years of age) were grouped with those who had received the vaccine early or on-time. Any factors with global significance (Prob>*F*) of *p*<0.1 in the univariable Poisson regressions were included in a multivariable Poisson regression model following a step-wise model selection strategy. Factors were kept in the final multivariable model if *p≤*0.05.

We conducted a sensitivity analysis to assess whether using Monday (first day in the assigned week of birth) or Sunday (last day in week in the assigned week of birth) as opposed to assigning a random day within the week, altered the conclusions made.

## RESULTS

Of the 1,782 children in this analysis, 919 (51.6%) were boys; the majority (97.2%) was from a white ethnic background. At the first MCS interview, 1,233 (69.2%) lived in ‘disadvantaged’ and 549 in ‘advantaged’ electoral wards, with 1,350 (75.8%) living in urban and 432 in rural areas. (Disadvantaged wards were oversampled (30), but the weights were adjusted in analyses making results nationally representative).

In total, 98.6% (*n*=1,759) received the first dose of DTP; with the majority 79.6% (*n=*1,429) receiving it between 8 and 12 weeks of age, 14.2% (*n=*251) early (before 8 weeks of age), 4.8% (*n=*79) late (over 12 weeks of age) and 1.4% (*n*=23) with no record of receipt by 14 years of age (18).

The number of children categorised as receiving DTP defined as late was 111, 64 and 79 when the day of birth was assigned to a Monday, Sunday or Thursday respectively. We explored associations with key variables using the different days and did not find any consistent differences in outcomes (data not shown). We chose to use the random day of birth for consistency with our earlier work and to reduce misclassification bias.

Delayed DTP vaccination (Table 2) was associated with characteristics of the child, mother and household. In the unadjusted analysis, delay was more likely for boys than girls (risk ratio 1.57 (95% CI 1.01, 2.44)), babies with older natural siblings (2.61 (1.55, 4.40)), those admitted to special or intensive care following delivery (2.24 (1.10, 4.57)) and babies whose birth weight was greater than 4Kg (1.95 (1.03, 3.69)). DTP vaccination was more likely to be delayed in children whose mothers had no academic qualifications (1.70 (1.04, 2.78)), were not in employment (2.25 (1.37, 3.67)), who smoked during the pregnancy (1.90 (1.08, 3.34)) and who did not breast feed (2.18 (1.27, 3.77)). Household factors found to be significantly associated with delayed DTP vaccination included a larger number of children in the household (for four or more children (4.55 (2.35, 8.81))), household income and income quintile, housing tenure and the mother’s partner not being in work (2.21 (1.15, 4.24)).

**Table 2.** Factors associated with late receipt of the first DTP vaccine

| Measure |  | Weighted % late (no.) | Unadjusted RR (95% CI) | p- value | Prob > F | Adjusted RR (95% CI) | p- value |
| --- | --- | --- | --- | --- | --- | --- | --- |
| Maternal age at child's birth - per 5 year increase |  |  | 0.78 (0.60 - 1.00) | 0.053 | 0.0525 | 0.73 (0.53 - 1.00) | 0.053 |
| Maternal academic qualifications | Degree | 1.2% (265) | 0.23 (0.07 - 0.72) | 0.012 | 0.0109 | 0.27 (0.08 - 0.90) | 0.034 |
|  | Diploma / A levels | 2.4% (309) | 0.46 (0.20 - 1.03) | 0.060 |  | 0.54 (0.21 - 1.43) | 0.21 |
|  | O level / GCSE | 5.3% (867) | 1 |  |  | 1 |  |
|  | Other | 3.6% (23) | 0.68 (0.12 - 4.02) | 0.67 |  | 0.60 (0.10 - 3.51) | 0.56 |
|  | None | 8.9% (316) | 1.70 (1.04-2.78) | 0.035 |  | 1.49 (0.90 - 2.46) | 0.12 |
| Maternal social class | Mangement & professional | 2.7% (467) | 1 |  | 0.0000 | 1 |  |
|  | Intermediate | 5.8% (283) | 2.17 (0.95 - 4.96) | 0.067 |  | 1.13 (0.48 - 2.65) | 0.77 |
|  | Small employer & self-employ | 0.0% (64) | n/a | <0.0001 |  | n/a | <0.0001 |
|  | Lower supervisory & technical | 2.4% (114) | 0.89 (0.23 - 3.46) | 0.86 |  | 0.30 (0.07 - 1.37) | 0.12 |
|  | Semi-routine & routine | 5.5% (707) | 2.07 (0.97 - 4.39) | 0.058 |  | 0.62 (0.28 - 1.38) | 0.24 |
|  | Never employed | 10.1% (125) | 3.79 (1.73 - 8.29) | 0.001 |  | 0.94 (0.35 - 2.49) | 0.89 |
| Maternal work status | In work / on leave | 3.0% (920) | 1 |  | 0.0016 |  |  |
|  | Not in work / on leave | 6.7% (862) | 2.25 (1.37 - 3.67) | 0.002 |  |  |  |
| Smoked during pregnancy | No or gave up | 3.9% (1295) | 1 |  | 0.0257 |  |  |
|  | Yes | 7.4% (487) | 1.90 (1.08 - 3.34) | 0.026 |  |  |  |
| Whether breastfed | Ever | 3.3% (1083) | 1 |  | 0.0056 |  |  |
|  | Never | 7.1 % (699) | 2.18 (1.27 - 3.77) | 0.006 |  |  |  |
| Sex | Female | 3.7% (863) | 1 |  | 0.0454 | 1 |  |
|  | Male | 5.8% (919) | 1.57 (1.01 - 2.44) | 0.045 |  | 1.53 (1.01 - 2.31) | 0.047 |
| Birth weight | <2.5Kg | 6.0% (90) | 1.44 (0.63 - 3.29) | 0.38 | 0.0725 | 0.83 (0.34 - 1.92) | 0.66 |
|  | 2.51 to 3 Kg | 3.7% (277) | 0.87 (0.39 - 1.95) | 0.74 |  | 0.91 (0.43 - 1.94) | 0.80 |
|  | 3.01 to 3.5Kg | 4.2% (693) | 1 |  |  | 1 |  |
|  | 3.51 to 4Kg | 4.8% (513) | 1.14 (0.55 - 2.36) | 0.73 |  | 1.14 (0.58 - 2.23) | 0.71 |
|  | >4 Kg | 8.2% (206) | 1.95 (1.03 - 3.69) | 0.040 |  | 2.02 (1.09 - 3.73) | 0.025 |
| Went to SCBU / ICU | No | 4.4% (1636) | 1 |  | 0.0271 | 1 |  |
|  | Yes | 9.8% (145) | 2.24 (1.10 - 4.57) | 0.027 |  | 3.15 (1.65 - 5.99) | 0.001 |
| Birth order | First born | 2.5% (751) | 1 |  | 0.0005 | 1 |  |
|  | Has older natural siblings | 6.6% (1031) | 2.61 (1.55 - 4.40) | <0.0001 |  | 3.82 (1.97 - 7.38) | <0.0001 |
| Total number of children in HH | 1 | 2.5% (748) | 1 |  | 0.0001 |  |  |
|  | 2 or 3 | 6.0% (921) | 2.37 (1.38 - 4.07) | 0.002 |  |  |  |
|  | 4 or more | 11.5% (113) | 4.55 (2.35 - 8.81) | <0.0001 |  |  |  |
| Household income per annum | £0.00 to <£10,400 | 6.2% (493) | 0.94 (0.56 - 1.55) | 0.79 | 0.0279 |  |  |
|  | £10,400 to <£20,800 | 6.6% (587) | 1 |  |  |  |  |
|  | £20,800 to <£31,200 | 1.2% (342) | 0.18 (0.06 - 0.50) | 0.001 |  |  |  |
|  | £31,200 to <£52,000 | 2.9% (224) | 0.44 (0.13 - 1.47) | 0.18 |  |  |  |
|  | £52,000 and above | 2.3% (60) | 0.35 (0.08 - 1.50) | 0.15 |  |  |  |
| Income weighted quintile | Lowest quintile (lowest income) | 6.9% (445) | 1 |  | 0.0007 | 1 |  |
|  | Second quintile | 6.2% (403) | 0.90 (0.49 - 1.65) | 0.72 |  | 1.07 (0.55 - 2.08) | 0.83 |
|  | Third quintile | 5.9% (355) | 0.86 (0.54 - 1.37) | 0.52 |  | 1.25 (0.70 - 2.23) | 0.45 |
|  | Fourth quintile | 0.5% (318) | 0.07 (0.02 - 0.22) | <0.0001 |  | 0.15 (0.04 - 0.51) | 0.003 |
|  | Highest quintile | 2.9% (261) | 0.43 (0.17 - 1.08) | 0.071 |  | 0.82 (0.33 - 2.06) | 0.68 |
| Housing tenure | Own or mortgage | 3.4% (1090) | 1 |  | 0.0133 |  |  |
|  | Rent | 7.2% (602) | 2.13 (1.24 - 3.65) | 0.007 |  |  |  |
|  | Living elsewhere | 5.1% (90) | 1.51 (0.56 - 4.07) | 0.41 |  |  |  |
| Whether partner is in work | In work / on leave | 4.0% (1255) | 1 |  | 0.0185 |  |  |
|  | Not in work / on leave | 8.8% (178) | 2.21 (1.15 - 4.24) | 0.019 |  |  |  |

RR - Risk Ratio
Missing Values: Maternal academic qualifications 2; Maternal social class 22; Birth weight 3; Admission to SCBU 1; Household income 76; Partner in work 349

In the final adjusted model (Table 2), children with older natural siblings were nearly four times as likely to have delayed DTP vaccination than those who were first born (risk ratio 3.82 (95% CI 1.97, 7.38)). Delay was more likely among children admitted as a baby to special/intensive care (3.15 (1.65, 5.99)), among boys (1.53 (1.01, 2.31) and among babies with a birth weight of over 4 Kg (2.02 (1.09, 3.73)). The risk of vaccine delay decreased with increasing maternal age (0.73 (0.53, 1.00) per 5-year increase) and maternal education to degree level (0.27 (0.08, 0.90)). Associations were also seen with specific categories of maternal social class and household income quintile, but there were no consistent trends.

## DISCUSSION

This study shows that whilst the majority of infants were vaccinated in a timely manner, delayed primary vaccination was more likely among children with older natural siblings, those admitted to special or intensive care following delivery, those whose birth weight was greater than 4Kg and boys. There was a reduced risk of delayed vaccination with increasing maternal age and for babies born to graduate mothers.

To our knowledge this is the first study linking data from a nationally representative cohort study with routine health records to explore factors associated with delayed primary vaccinations in a UK setting. The high consent rate to link vaccination records, which provide accurate vaccination histories, with the rich data in the MCS enabled us to explore associations between vaccine delay and a range of socio-demographic factors. However, as the study was based on Welsh participants in the MCS who had not been interviewed outside Wales at any of the MCS sweeps, we were unable to explore potential associations with UK country or residential mobility in and out of Wales. We were also unable to explore associations with maternal and child ethnicity, first language and language spoken at home because of small numbers from minority ethnic groups in the study population. At present due to lack of a unified child health database in England, it is not possible to easily replicate this analysis for the UK as a whole.

We did not have access to the children’s actual dates of birth to calculate ages at vaccination. However, using the random day within the week of birth should not have affected the proportions reported as early, on-time or delayed (32). Ideally children should have received the first dose of the DTP vaccine as close to 8 weeks of age as possible. In the absence of an official definition of delay, we chose to define it according to the time the second vaccine was due. Although, using the random day of birth will have resulted in some misclassification of those who were and were not delayed based around the 12 weeks of age cut-off point, the random nature of this misclassification, should have prevented bias in our findings but may have weakened some of the associations reported for factors associated with vaccination delay. Furthermore, we tested alternative strategies for assigning day of birth in a sensitivity analysis and these did not materially affect our findings.

The majority of children in this cohort were immunised on time. However, consistent with other studies from many countries, children with older siblings were nearly four times as likely to have delayed DTP vaccination, than first born children (7, 9). This may reflect challenges accessing services as a result of competing demands on parents’ time, making attending appointments difficult or decreased parental worry (33). Babies who had been admitted to a special or intensive care following delivery were more than three times likely to be delayed receiving their first DTP vaccination. We did not have data on babies’ length of stay in special care units, but some may have been hospitalised for significant periods of time and opportunities for timely vaccination in hospital may have been missed. Babies admitted to special care units are likely to have conditions that make them not only more likely to catch a vaccine preventable disease, but to be more vulnerable to the effects of the disease and so are in particular need of protection by vaccination. Vaccination may be delayed because health professionals or parents incorrectly believe vaccination is contraindicated (34). On discharge, even after a short hospital stay, parents of these children experience stress which may persist (35) and once home, over-protective parents may delay attending routine appointments including for vaccination; vaccinations may not be prioritised in discharge plans and parents may delay vaccinations so their child can ‘have a rest’ after discharge (36). In contrast with some studies, we did not find an association between prematurity and vaccination delay (34); indeed a review by Sisson et al, shows a lack of consensus on effects of hospitalisation on vaccination timeliness for preterm or low birth weight infants (37). Although studies have shown that hospitals do not always use opportunities to immunise older children (38, 39), hospital based immunisation initiatives have been shown to be effective in ensuring preterm babies are immunised on time (40). We are unable to explain our finding that boys were more likely to have delayed vaccination than girls, although similar unexplained associations have been found in other studies. In a 1993 study in Liverpool, parents of boys were less likely to consent to pertussis vaccine than parents of girls (41). More recently and also using MCS data, Pearce et al found that girls were significantly less likely to be unimmunised against MMR at age three years, than boys (29). Both these observations were made in the years following vaccine safety controversies. Our finding that heavier babies were more likely to have delayed vaccination is unexplained, although as higher birth weight is associated with complicated deliveries and conditions notably gestational diabetes, delayed attendance for vaccination may result from poorer maternal health status.

Consistent with other studies, the risk of vaccine delay decreased with increasing maternal age (22, 42) and delay was associated with lower maternal academic qualifications (8). Associations were seen with specific categories of maternal social class and household income quintile, but there was no consistent trend with these factors as a whole. Other studies (13, 43, 44) using either the Welsh or Scottish Index of Multiple Deprivation have found that associations between deprivation and vaccination uptake and timeliness increase with the age of the child and number of vaccine doses.

Our analysis was not based on a current cohort of children and hence the estimates of delayed vaccination may not reflect the current picture. Similarly, our focus was on DTP vaccine, which is now given as part of a hexavalent vaccine DTaP/IPV/Hib/HepB (6-in-1) introduced in UK in 2017. In 2009 NICE published guidance on improving vaccine uptake (45). Although we have described the characteristics of the children in whom immunisation was delayed and of their families, we are unable to explore whether the reasons for delay were intentional or not. In the USA, Smith found that 21.8% of parents reported intentionally delaying vaccinations for their children (aged 19 to 35 months). Of these, 44.8% did so because of concerns about vaccine safety or efficacy and 36.1% delayed because of an ill child (46). However more recently Homel et al (11) reported parental attitudes contributed a relatively small percentage of delay. Our findings would suggest that in this cohort, delay arose from the complexities of life arising from social disadvantage, busy lives with large families and babies requiring special care and this is borne out by other studies. Recently there has been intense focus on the issue of ‘vaccine hesitancy’ a term used to refer to a “delay in acceptance or refusal of vaccination despite availability of vaccination services” (47) with suggestions that vaccine hesitancy is increasing, however, since data are not routinely gathered on timeliness of vaccination, it is not possible to comment on whether delay in vaccination, one element of vaccine hesitancy, has increased.

We consider that socio-demographic factors associated with delay in vaccination are unlikely to have altered significantly and these findings provide insights into where efforts to improve vaccine timeliness should be best targeted. To monitor this in future we would suggest routine assessment of vaccination timeliness.

Delayed receipt of the first dose of primary immunisations has been found to predict subsequent incomplete immunisation. Therefore, improving the timeliness of the first dose may improve both timeliness of subsequent doses and of completed courses of vaccination and thus vaccine uptake rates. This study assists in identifying those groups of infants who are at increased risk of delayed vaccination. In the absence of national data on vaccination timeliness, awareness of factors associated with delay may allow targeted interventions for those at highest risk. For infants admitted to a special or intensive care this could include offering vaccination prior to discharge or ensuring that appropriate communication takes place with their General Practitioner for vaccination in the community. Provision of clear, easy to read information, with flexible and easy to access services may facilitate vaccination of infants growing up in large families or where there are competing demands on parents’ time. Opportunistic catch-up during routine contact with health professionals would also reduce the inequalities that exist. More widely, this study underlines the importance of maternal education in facilitating positive health outcomes for children.

## Conclusions

This study demonstrates the benefits of using rich cohort data linked to routine child health data and to our knowledge, is the first study exploring socio-demographic factors associated with delayed receipt of primary immunisations, within a UK setting. Timely immunisation is important to provide children with maximum protection against serious infectious diseases.

## Supporting information

COI disclosure

STROBE checklist

## Data Availability

All data produced in the present work are contained in the manuscript.

## Acknowledgements

The authors are grateful to the Centre for Longitudinal Studies, UCL Institute of Education and the UK Data Service as well as the child health offices who enter data and NHS Wales Digital Health and Care Wales (DHCW), formerly known as the NHS Wales Informatics Service (NWIS) that maintain the CCH2000 CHIS system and central data record, who provide anonymised data held in the Secure Anonymised Information Linkage (SAIL) Databank, which is part of the national e-health records research infrastructure for Wales. This study makes use of anonymised data held in the Secure Anonymised Information Linkage (SAIL) Databank. We would like to acknowledge all the data providers who make anonymised data available for research. All research conducted has been completed under the permission and approval of the SAIL independent Information Governance Review Panel (IGRP) project number 0410.

## Authors’ contributions

HB and SW conceived and designed the study. KT, AB, LG, SW and AA were responsible for data curation. SW carried out the data analysis with support from HB and MC-B. SW and HB wrote the manuscript and all authors contributed to critically appraising and review of the manuscript. All authors approved the final manuscript.

