## Supplementary material for "Linking cohort data and Welsh routine health records to investigate children at risk of delayed primary vaccination": COI disclosure

### ICMJE DISCLOSURE FORM

| | |
|---|---|
| **Date:** | 3/13/2022 |
| **Your Name:** | Suzanne Walton (on behalf of all authors) |
| **Manuscript Title:** | **Linking cohort data and Welsh routine health records to investigate children at risk of delayed primary vaccination** |
| **Manuscript Number (if known):** | Click or tap here to enter text. |

In the interest of transparency, we ask you to disclose all relationships/activities/interests listed below that are related to the content of your manuscript. "Related" means any relation with for-profit or not-for-profit third parties whose interests may be affected by the content of the manuscript. Disclosure represents a commitment to transparency and does not necessarily indicate a bias. If you are in doubt about whether to list a relationship/activity/interest, it is preferable that you do so.

The author's relationships/activities/interests should be defined broadly. For example, if your manuscript pertains to the epidemiology of hypertension, you should declare all relationships with manufacturers of antihypertensive medication, even if that medication is not mentioned in the manuscript.

In item #1 below, report all support for the work reported in this manuscript without time limit. For all other items, the time frame for disclosure is the past 36 months.

| | | **Name all entities with whom you have this relationship or indicate none (add rows as needed)** | **Specifications/Comments (e.g., if payments were made to you or to your institution)** |
|---|---|---|---|
| | | Time frame: Since the initial planning of the work | |
| 1 | All support for the present manuscript (e.g., funding, provision of study materials, medical writing, article processing charges, etc.) **No time limit for this item.** | ☐ None | |
| | | This work was supported by the Wellcome Trust (grant number 087389/B/08/Z) | All authors apart from RR |
| | | Supported by Health Data Research UK (HDR-9006), which is funded by the UK Medical Research Council, Engineering and Physical Sciences Research Council, Economic and Social Research Council, National Institute for Health Research (England), Chief Scientist Office of the Scottish Government Health and Social Care Directorates, Health and Social Care Research and Development Division (Welsh Government), Public Health Agency (Northern Ireland), British Heart Foundation and Wellcome. | LG, CD, RAL and AA |
| | | Funded by the Asthma UK Centre for Applied Research (AUKAC- 2012-01) | RAL |
| | | Supported by an ESRC award establishing the Administrative Data Research Centre Wales (ES/L007444/1) | KT |
| | | Supported by the ADR Wales programme of work. The ADR Wales programme of work is aligned to the priority themes as identified in the Welsh Government's national strategy: Prosperity for All. ADR Wales brings together data science experts at Swansea University Medical School, staff from the Wales Institute of Social and Economic Research, Data and Methods (WISERD) at Cardiff University | AA, AB, LG and RAL |

| | | Name all entities with whom you have this relationship or indicate none (add rows as needed) | Specifications/Comments (e.g., if payments were made to you or to your institution) |
|---|---|---|---|
| | | and specialist teams within the Welsh Government to develop new evidence which supports Prosperity for All by using the SAIL Databank at Swansea University, to link and analyse anonymised data. ADR Wales is part of the Economic and Social Research Council (part of UK Research and Innovation) funded ADR UK (grant ES/S007393/1). | |
| | | Supported by Barts Charity (Grant/Award Number: MGU0419). | CD |
| | | Supported by the National Centre for Population Health and Well-Being Research (NCPHWR) which is funded by Health and Care Research Wales. | AB |
| | | The Millennium Cohort Study [http://dx.doi.org/10.14301/llcs.v7i4.410] is funded by grants to the Centre for Longitudinal Studies at the Institute of Education from the Economic and Social Research Council and a consortium of government departments. | |
| | | Research at the UCL Institute of Child Health and Great Ormond Street Hospital for Children receives a proportion of the funding from the Department of Health's National Institute for Health Research Biomedical Research Centres funding scheme. | SW, MCB, LJG, HB |
| **Time frame: past 36 months** | | | |
| **2** | Grants or contracts from any entity (if not indicated in item #1 above). | ☐ **None** | |
| | | UKRI – Medical Research Council | RAL<br>Health and Care Research Wales |
| | | UKRI-Economic and Social Research Council | RAL |
| | | Office for National Statistics funding for KT from Feb 2019-present. | KT |
| | | HDR-UK grant "Ethnicity and COVID-19: investigating the determinants of excess risk" | KT |
| **3** | Royalties or licenses | ☒ **None** | |
| **4** | Consulting fees | ☒ **None** | |
| **5** | Payment or honoraria for | ☒ **None** | |

| | | Name all entities with whom you have this relationship or indicate none (add rows as needed) | Specifications/Comments (e.g., if payments were made to you or to your institution) |
|---|---|---|---|
| | lectures, presentations, speakers bureaus, manuscript writing or educational events | | |
| 6 | Payment for expert testimony | ☒ None | |
| 7 | Support for attending meetings and/or travel | ☒ None | |
| 8 | Patents planned, issued or pending | ☒ None | |
| 9 | Participation on a Data Safety Monitoring Board or Advisory Board | ☒ None | |
| 10 | Leadership or fiduciary role in other board, society, committee or advocacy group, paid or unpaid | ☐ None | |
| | | Member, Board of Trustees , Play Wales, a charity | RAL |
| | | Member of the Social Statistics Committee for the Royal Statistical Society | KT |
| | | Senior Statistical Advisory for Buckinghamshire Disability Service | KT |
| 11 | Stock or stock options | ☒ None | |
| 12 | Receipt of equipment, materials, drugs, medical writing, | ☒ None | |

| | | Name all entities with whom you have this relationship or indicate none (add rows as needed) | Specifications/Comments (e.g., if payments were made to you or to your institution) |
|---|---|---|---|
| | gifts or other services | | |
| **13** | Other financial or non-financial interests | ☒ **None** | |

**Please place an "X" next to the following statement to indicate your agreement:**

☒ I certify that I have answered every question and have not altered the wording of any of the questions on this form.
